# Clinical Impact of Multiplex Molecular Diagnostic Testing in Children with Acute Gastroenteritis Presenting to An Emergency Department: A Multicenter Prospective Study

**DOI:** 10.1101/2023.07.27.23293208

**Authors:** Andrew T. Pavia, Daniel M. Cohen, Amy L. Leber, Judy A. Daly, Jami T. Jackson, Rangaraj Selvarangan, Neena Kanwar, Jeffrey M. Bender, Jennifer Dien Bard, Ara Festekjian, Susan Duffy, Chari Larsen, Kristen M. Holmberg, Tyler Bardsley, Benjamin Haaland, Kevin M. Bourzac, Christopher Stockmann, Kimberle C. Chapin, Daniel T. Leung

**Affiliations:** Departments of Pediatrics and Internal Medicine, Spencer Fox Eccles School of Medicine, University of Utah, Salt Lake City, UT, USA; Department of Pediatrics, Nationwide Children’s Hospital, Columbus, OH, USA; Department of Pathology, Spencer Fox Eccles School of Medicine, University of Utah, Salt Lake City, UT, USA; Children’s Mercy Hospital, Kansas City, MO, USA; Children’s Hospital of Los Angeles; Keck School of Medicine, University of Southern California, Los Angeles, CA, USA; bioMérieux, Salt Lake City, UT, USA; Department of Population Health Sciences, University of Utah School of Medicine, Salt Lake City, UT, USA; Department of Pathology and Laboratory Medicine, Rhode Island Hospital, Alpert Medical School, Brown University, Providence, RI, USA; Department of Emergency Medicine, Hasbro Children’s Hospital, Alpert Medical School, Brown University, Providence, RI, USA

**Keywords:** Gastroenteritis, diarrhea, multiplex panels, diagnosis, outcomes, pediatric, polymerase chain reaction

## Abstract

**Background:** Multiplex molecular diagnostic panels have greatly enhanced detection of gastrointestinal pathogens. However, data on the impact of these tests on clinical and patient-centered outcomes are limited.

**Methods:** We conducted a prospective, multicenter, stepped-wedge trial to determine the impact of multiplex molecular testing at five academic children’s hospitals in children presenting to the ED with acute gastroenteritis. Caregivers were interviewed on enrollment and again 7-10 days after enrollment to determine symptoms, risk factors, subsequent medical visits, and impact on family members. During the pre-intervention period, diagnostic testing was performed at the discretion of clinicians. During the intervention period, multiplex molecular testing was performed on all children with results available to clinicians. Primary outcome was return visits to a health care provider within 10 days of enrollment.

**Results:** Potential pathogens were identified by clinician ordered tests in 19/571 (3.3%) in the pre-intervention period compared to 434/586 (74%) in the intervention period; clinically relevant pathogens were detected in 2.1% and 15% respectively. In the multivariate model adjusting for potential confounders, the intervention was associated with a 21% reduction in the odds of any return visit (OR 0.79; 95% CI 0.70-0.90). Appropriate treatment was prescribed in 11.3% compared to 19.6% during the intervention period(P=0.22).

**Conclusions:** Routine molecular multiplex testing for all children presenting to the ED with AGE detected more clinically relevant pathogens and led to a 21% decrease in return visits. Additional research is needed to define patients most likely to benefit from testing.

## INTRODUCTION

Acute gastroenteritis is a major cause of emergency department (ED) visits and hospitalization in countries with advanced economies and a major cause of death and disability worldwide.[1, 2] For many children with gastroenteritis, rehydration, antiemetics, and reassurance are the only therapy needed. However, a range of bacterial and protozoal infections may require definitive diagnosis and targeted therapy.[3, 4] Multiplex molecular diagnostic panels available in recent years greatly enhance detection of a wide range of pathogens.[5-10] The potential benefits of rapid and accurate detection include decreased health care utilization, decreased parental anxiety, appropriate antimicrobial therapy, and improved infection control and outbreak recognition.[11] The potential negative consequences include overtreatment of incidentally detected organisms, increased cost, and the potential failure to provide isolates to public health laboratories for molecular characterization.[12]

Relatively few studies to date have prospectively evaluated the impact of multiplex molecular testing on improving clinical and patient-centered outcomes. We conducted a multicenter prospective pre-post intervention study to evaluate the direct and indirect impact of the introduction of a multiplex molecular diagnostic panel on care for children presenting to pediatric Emergency Departments (EDs) with acute gastroenteritis.

## METHODS

### Design

The objective of the FilmArray GI Panel IMPACT (**I**mplementation of a **M**olecular Diagnostic for **P**ediatric **Ac**u**t**e Gastroenteritis) study was to determine the clinical impact of multiplex molecular diagnostic testing for stool pathogens versus clinician-selected diagnostic testing in children presenting to EDs with acute gastroenteritis. We conducted a prospective, multicenter pragmatic-stepped wedge study at five academic children’s hospitals. (**Supplemental Figure 1**) Children were enrolled between April 2015 and September 2016. Enrollment was staggered in a stepped-wedge fashion with the goal of reducing the potential impact of seasonal variation on the etiology of diarrhea. The start of the study, however, was impacted based on each institution’s timeline for completing study. The study was approved by the institutional review board at each institution.

### Eligibility

Children <18 years presenting to the ED or on-site urgent care center were eligible if they had symptoms of gastroenteritis (diarrhea or nausea/vomiting as a chief complaint) for at least 24 hours. Children were excluded if symptoms were present <24 hours or ≥14 days, if a diagnosis other than gastroenteritis was apparent (e.g. appendicitis, inflammatory bowel disease), if they or a family member had been previously enrolled in the study, or if informed consent could not be obtained.

### Study procedures and intervention

Written informed consent was obtained from parents or legal guardians; children provided assent as age-appropriate. Trained study personnel interviewed subjects using a standardized questionnaire at enrollment. The questionnaire included demographic information, symptoms, epidemiologic exposures and previous treatment. Study personnel interviewed subjects again by telephone 7-10 days after enrollment to determine duration of illness, subsequent medical visits, tests or treatment, secondary illness in the family, and impact on family members. Medical charts were systematically abstracted after the final contact to collect laboratory tests ordered, results, treatments prescribed, hospital admission and return visits.

All children were asked to provide a stool specimen during the visit or within 48 hours for multiplex PCR testing, regardless of whether the testing was ordered for clinical purposes. Specimens were placed in Cary-Blair transport medium. During the pre-intervention period, diagnostic testing was at the discretion of the clinicians in the ED. Stool specimens were stored and later tested by multiplex PCR, but results were not available to the clinicians or patients. After roughly 100 evaluable patients were enrolled in the pre-intervention period, each site prepared to transition to the intervention period.

Before initiating the intervention period, clinicians in each ED were educated about the multiplex PCR panel, including performance characteristics, clinical features of each pathogen, recommended management, and explanations in patient-friendly language (http://cstockmann.github.io/IMPACT/index.html). Stool samples from children enrolled in the intervention period were tested by multiplex PCR in real-time at the hospital laboratory (with results generally available within ∼1-4 hours of receipt) and communicated to the ED. If the patient had been discharged before results were available, clinicians contacted the family by the next morning. Clinicians were free to order additional testing at their discretion.

During the pre-intervention period, children were included in the analysis regardless of whether a specimen was provided for multiplex PCR; during the intervention period, children were excluded if they could not provide a stool specimen within 48 hours of enrollment.

### Laboratory testing

Clinician-selected testing was performed in the hospital laboratory by standard methods and included stool culture, *Clostridiodes difficile* testing by EIA or nucleic acid amplification, rotavirus and adenovirus antigen detection by EIA, norovirus detection by nucleic acid amplification, or protozoal detection by microscopy or EIA for *Giardia lamblia/duodenalis* and *Cryptosporidium*. We used the BIOFIRE FILMARRAY Gastrointestinal (GI) Panel assay for multiplex PCR testing (bioMérieux, Salt Lake City, UT). This platform detects 22 pathogens. **(Supplemental Table 1**) [5] During the intervention period, when pathogens of public health significance were detected by PCR (*Campylobacter spp., Salmonella spp.*, Shiga toxin-producing *Escherichia coli* [STEC],STEC, *Shigella spp.*/EIEC), specimens were reflexed to culture and/or sent to Public Health laboratories.

### Outcome measures

The primary outcome was return visits to a health care provider within 10 days of enrollment. Secondary outcomes included number of return visits, return visit to ED or hospitalization, number of pathogens detected, number of potentially treatable pathogens detected (*Campylobacter, Shigella, Vibrio sp., Yersinia*, *Plesiomonas*, *Cryptosporidium, Cyclospora cayetanensis, Entamoeba histolytica* and *G. lamblia*), clinically relevant pathogens detected – e.g. those that might alter care (potentially treatable pathogens plus those for which withholding antibiotics is important, i.e. *Salmonella* and STEC), and the proportion of children receiving appropriate treatment. Since the clinical significance of *C. difficile* detection by PCR alone in outpatients is challenging to interpret, we reported total detections and detection of *C. difficile* as the sole pathogen in children >2 years of age but did not define it as a potentially treatable pathogen. We also measured the impact on secondary illness in family members, absence from daycare, school, and work, parental response to knowing the diagnosis, and detection of outbreaks.

### Statistical analysis

Power calculations were based on historical data on return visits for patients with gastroenteritis seen at Primary Children’s Hospital showing a mean of 0.5 additional healthcare encounters per patient within seven days of the initial encounter. Sample size was limited by funding to 1100, providing 70% power to detect a 25% reduction in return visits with an alpha of 0.05.

Patient characteristics, clinical findings, tests ordered, pathogens detected, and outcomes were summarized using descriptive statistics for pre and post-intervention periods and compared using chi-squared or Mann-Whitney Wilcoxon tests for categorical or continuous data. Differences were presented as odds ratios and exact confidence intervals. To account for imbalances in the study periods, the primary analyses of the impact of multiplex molecular diagnostic testing on return visits (any return, ED return or hospitalization, and number of return visits within 10 days of enrollment) was based on logistic or negative binomial generalized estimating equations models, as appropriate, accounting for within site clustering, and adjusted for patient age, insurance, illness duration, pathogen type, gender, race, ethnicity, diarrhea, fever, constipation, number of stools in last 24 hours, and quarter by year.[13, 14] The consistency of the estimated impact of multiplex molecular diagnostic testing across subpopulations of interest defined by site, gender, race, patient age, and illness duration was assessed via subgroup analyses mirroring the full analysis. To assess the impact of a *Shigella* outbreak in Kansas City during the time of study, we conducted sensitivity analyses omitting Children’s Mercy Hospital (CMH) observations. [15]

## RESULTS

### Demographics

Between April 2015 and September 2016, we enrolled 1,157 patients (571 in the pre-intervention period and 586 in the intervention period). **(Figure 1)** The mean age was 4.9 years; 64% were under 5 years of age. **(Table 1)** Race/ethnicity and season of enrollment differed between the pre-intervention and intervention periods as a result of the staggered enrollment by site. **(Table 1 and supplemental Figure 1).** The majority of children presented with vomiting and diarrhea; diarrhea was bloody in 13% **(Table 2).** More children in the intervention period had diarrhea and more had fever in the pre-intervention period. Symptoms were present for a median of 2 days (IQR: 1-5) before presentation.

**Figure 1:**
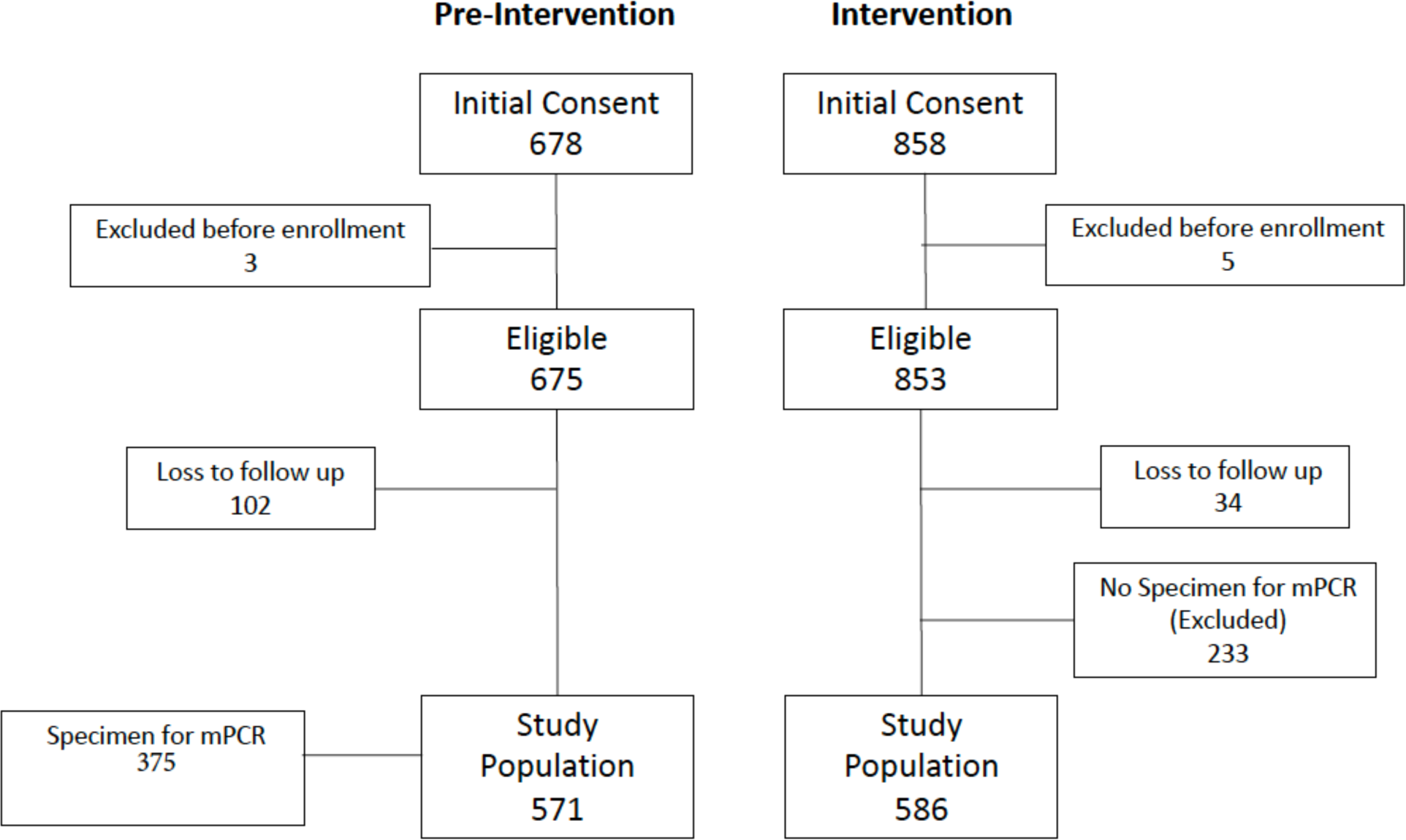
Consort diagram of subject disposition

**Table 1:** Demographic and exposure characteristics by study period.

| **Patient characteristics** | **Pre-intervention**<br>**N =571** | **Post-intervention**<br>**N =586** | **p-value** |
| --- | --- | --- | --- |
| **Female gender** | 300 (52.5%) | 272 (46.4%) | 0.037 |
| **Race** |  |  | <0.001 |
| White | 237 (42%) | 343 (59%) |  |
| Black | 149 (26%) | 135 (23%) |  |
| Other/Unknown | 185 (32%) | 108 (18%) |  |
| **Ethnicity** |  |  | <0.001 |
| Hispanic/Latino | 181 (32 %) | 167 (29%) |  |
| Non-Hispanic or Latino | 363 (64%) | 412 (70%) |  |
| Not specified/refused | 27 (5%) | 7 (1%) |  |
| **Age (Mean/Median)** | 4.9 (3) | 4.9 (3) | 0.94 |
| **Age Group** |  |  | 0.15 |
| <6 months | 53 (9.3%) | 63 (11%) |  |
| 6-23 months | 164 (29%) | 83 (31%) |  |
| 2-4 years | 155 (27%) | 122 (21%) |  |
| 5-11 years | 132 (23%) | 150 (12%) |  |
| 12-17 years | 67 (12%) | 68 (12%) |  |
| **Underlying Medical Condition** | 144 (25%) | 158 (27%) | 0.50 |
| **Immunosuppressed** | 3 (2%) | 6 (4%) | 0.51 |
| **Insurance** |  |  | 0.46 |
| Public | 375 (66%) | 368 (63%) |  |
| Private | 170 (30%) | 194 (33%) |  |
| None/Other | 26 (5%) | 24 (4%) |  |
| **Attend pre-school or daycare** | 85 (32.8%) | 72 (28.7%) | 0.20 |
| **Pet or animal exposure** | 320 (65%) | 331 (65%) | 0.97 |
| **International travel (past month)** | 9 (2%) | 14 (3%) | 0.33 |
| **Season** |  |  | <0.001 |
| Summer (Jul-Sep) | 310 (54%) | 66 (11%) |  |
| Fall (Oct-Dec) | 94 (16%) | 202 (34%) |  |
| Winter (Jan-Mar) | 45 (8%) | 214 (37%) |  |
| Spring (Apr-Jun) | 122 (21%) | 104 (18%) |  |

**Table 2:** Clinical features at presentation, and tests ordered by study period.

|  | Pre-Intervention<br>N= 571 | Post-intervention<br>N=586 | P value |
| --- | --- | --- | --- |
| Fever | 324 (57%) | 291 (50%) | 0.0123 |
| Vomiting | 450 (79%) | 459 (78%) | 0.98 |
| Vomiting duration in days<br>before enrollment<br>(median [IQR]) | 2 (1-3) | 2 (1-3) | 0.29 |
| Diarrhea | 473 (83%) | 547 (93%) | <0.0001 |
| Diarrhea duration in days<br>before enrollment<br>(median [IQR]) | 2 (1-5) | 3 (2-5) | 0.077 |
| Median stools/day | 4 (1-7) | 5 (2-8) | <0.0001 |
| Bloody diarrhea | 52 (11%) | 74 (13%) | 0.25 |
| Antibiotics prescribed<br>before ED visit | 20 (3.5%) | 13 (2.2%) | 0.19 |
| Antibiotics prescribed<br>during ED stay | 19 (3%) | 24 (4%) | 0.40 |
| Admitted to hospital | 81 (14%) | 96 (16%) | 0.30 |
| **Diagnostic testing** |  |  |  |
| Stool culture ordered | 70 (12%) | 85 (14%) | 0.26 |
| STEC study ordered | 33 (6%) | 44 (8%) | 0.24 |
| Viral studies ordered | 10 (2%) | 15 (3%) | 0.34 |
| *C. difficile* studies ordered | 28 (5%) | 30 (5%) | 0.87 |
| Ova and parasite exam<br>ordered | 17 (3%) | 24 (4%) | 0.30 |
| *Giardia/Cryptosporidia*<br>DFA ordered | 15 (3%) | 17 (3%) | 0.78 |

### Microbiology

During the pre-intervention period, clinicians ordered etiologic testing on 80 (14%) of 571 patients. **(Table 2).** Diagnostic tests were more likely to be ordered if the stool was reported as bloody (33/52 [64%]) than non-bloody (43/402 [11%]; Odds Ratio [OR] 14.0, 95% Confidence Intervals [CI] 7.6-27.7). Testing was not associated with reported fever; tests were obtained in 40/324 (12.4%) children with fever and 38/239 (15.9%) without fever (OR 0.75; 95% CI 0.46-1.20).

Clinician-ordered testing detected a pathogen in 19 (3.3%) of 571 patients during the pre-intervention period, including 14 (2.5%) with a treatable pathogen and 16 (2.8%) with a clinically relevant pathogen. **(Table 3)** Stool was available for batched testing by multiplex testing in 375/571 patients during the pre-intervention period. A total of 383 potential pathogens were detected in 262 patients (70%). Viral pathogens were detected in 135 (36%); EPEC, norovirus, *C. difficile*, and *Shigella*/EIEC were the most commonly detected pathogens overall. *C. difficile* was detected in 43 (11.5%) of 375 children by molecular testing, however, *C. difficile* was the sole pathogen in only 8 children ≥2 years of age.

**Table 3.** Pathogens detected by clinician-ordered standard of care tests and by multiplex PCR by study period. During Pre-intervention period, multiplex PCR testing was performed on stored stool specimens and not available to clinicians. During Intervention period multiplex testing was performed in real-time and available to clinicians.

|  | Pre-Intervention |  | Intervention |  |
| --- | --- | --- | --- | --- |
|  | Standard of care clinician-ordered tests (N=571) | Multiplex PCR (N=375)<br>(Clinician blinded to results) | Standard of care clinician-ordered tests (N=586) | Multiplex PCR (N=586)<br>(Results available to clinician) |
| **Bacteria** |  |  |  |  |
| *Campylobacter* | 4 (0.7%) | 13 (3.5%) | 1 (0.2%) | 11 (1.9%) |
| *Salmonella* | 2 (0.4%) | 11 (2.9%) | 6 (1.0%) | 18 (3.1%) |
| *Shigella/EIEC* | 9 (1.6%) | 33 (8.8%) | 4 (0.8%) | 24 (4.1%)* |
| *Plesiomonas* |  | 0 (0) |  | 2 (0.3%) |
| *Yersinia* |  | 0 (0) |  | 2 (0.3%) |
| STEC | 4 (0.7%) | 14 (3.7%) | 1 (0.2%) | 14 (2.4%) |
| *E. coli* O157 | 4 (0.7%) | 3 (0.8%) | 1 (0.2%) | 3 (0.5%) |
| ETEC |  | 10 (2.7%) |  | 6 (1.0%) |
| EAEC |  | 21 (5.6%) |  | 36 (6.1%) |
| EPEC |  | 76 (20%) |  | 67 (11.4%) |
| *C. difficile* | 2 (0.4%) | 43 (11.5%) | 6 (0.6%) | 94 (16.0%)† |
| *C. difficile* no virus and age ≥ 2 years | 1 (0.2%) | 8 (2.1%) | 1 (0.2%) | 23 (3.9) |
| **Viruses** |  |  |  |  |
| Adenovirus F 40/41 | 1 (0.2%) | 33 (8.8%) | 1 (0.2%) | 61 (10.4%) |
| Astrovirus |  | 6 (1.6%) |  | 43 (7.3%)* |
| Norovirus GI/GII |  | 57 (15.2%) |  | 148 (25.3%)* |
| Rotavirus | 2 (0.4%) | 16 (4.3%) | 1 (0.2%) | 12 (2.0%) |
| Sapovirus |  | 31 (8.3%) |  | 66 (11.3%) |
| Any viral pathogen | 3 (0.6%) | 135 (36%) | 2 (0.4%) | 294 (50%)* |
| **Protozoa** |  | 18 (3.1) |  | 23 (3.9%) |
| *Cryptosporidium* |  | 10 (2.7%) |  | 14 (2.4%) |
| *Cyclospora* |  | 0 (0) |  | 0 (0) |
| *Giardia* |  | 9 (2.4%) |  | 9 (1.5%) |
| At least 1 potential pathogen | 19 (3.3%) | 262 (70%) | 15 (3%) | 434 (74%) |
| Any treatable pathogen ** | 14 (2.5%) | 65 (17.3%) | 5 (0.9%) | 61 (10.4%) |
| Any clinically relevant pathogen†† | 16 (2.8%) | 84 (22.4%) | 12 (2%) | 88 (15%) |

\* P <0.01 for difference between pre-intervention and intervention prevalence by multiplex PCR
† P<0.05 for difference between pre-intervention and intervention prevalence by multiplex PCR
\*\* Defined as: *Campylobacter*, *Shigella*, *Vibrio sp.*, *Yersinia*, *Plesiomonas*, *Cryptosporidium*, *Cyclospora cayetanensis*, *Entamoeba histolytica* and *G. lamblia*.
†† Defined as *Campylobacter*, *Shigella*, *Vibrio sp.*, *Yersinia*, *Plesiomonas*, *Cryptosporidium*, *Cyclospora cayetanensis*, *Entamoeba histolytica*, *G. lamblia*, *Salmonella sp.*, and *STEC*.

During the pre-intervention period, potentially treatable pathogens were detected by multiplex molecular testing in 65 (17.3%) of 375 patients tested compared to 12 (2.1%) of 571 children by clinician ordered testing (difference 14.3%, 95% CI 10.3%-18.2%; P<0.001). Clinically relevant pathogens were detected in 84 (22.4%) by multiplex molecular testing compared to 14 (2.5%) by clinician-ordered testing (difference 18.6%; 95% CI 14.1%-23.0%; P<0.001).

During the intervention period, 627 potential pathogens were detected by multiplex molecular tesing in 434 (74%) of 571 children. Norovirus, *C. difficile*, EPEC and sapovirus were the most common pathogens detected. Only 23 children (3.9%) with *C. difficile* were ≥2 years of age and had no other pathogens detected. Viral pathogens were detected in 294 patients (50%) during the intervention period, significantly more than during the pre-intervention period (36%; P<0.001). *Shigella spp.*/EIEC was detected less frequently during the intervention period (24 [4.1%] vs 33 [8.8%]; P=0.003). This was driven by a community-wide outbreak of *Shigella* in Kansas City impacting CMH during the pre-intervention period. Multiplex PCR results were available and shared with the family before leaving the ED in 141 (24%) cases.

### Outcomes

The likelihood of follow-up visits was not significantly lower during the intervention period in univariate analysis. During the intervention period 186 (32%) children had at least one follow-up visit with a health care provider after the enrollment visit to the ED compared to 174 (30%) during the pre-intervention period (OR 1.06 95% CI 0.82-1.37). Of these 54 (9.2%) were hospitalized or returned to urgent care or the ED during the intervention period, compared to 53 children (9.3%) during the pre-intervention period (OR 1.00; 95% CI 0.66-1.53).

During intervention period, children with a viral pathogen detected were less likely to have a return visit (75 of 294 [26%]) compared to those without a virus detected (111 0f 292 [38%], OR 0.56; 95% CI 0.39-0.79; P = 0.001). This was not the case during the pre-intervention period (39 of 135 [29%] vs 72 of 240 [30%]; OR 0.95 95% CI 0.60-1.51). During the intervention period children who were enrolled in winter were less likely to have a repeat visit (62 of 217 [29%] compared to 21 of 44 [48%]; OR 0.44; 95% CI 0.23-.85; P= 0.01) during the pre-intervention period. Conversely, during the intervention period those enrolled during summer were more likely to have a return visit (28 of 66 [42%] vs. 86 of 305 [28%]; OR 1.9; 95% CI 1.08-3.2; P = 0.023).

In the multivariate model adjusting for age, insurance, illness duration, pathogen type, sex, race, ethnicity, diarrhea, fever, number of stools, and quarter by year of patient presentation, the intervention was associated with a 21% reduction in the odds of any return visit (OR 0.79; 95% CI 0.70-0.90; P=<0.001; Table 4). Multivariate results for the number of return visits were similar (rate ratio 0.83; 95% CI 0.70-0.97; P=0.023; Table 4). However, multivariate analyses did not show a statistically significant difference between pre- and post-intervention periods in ED visits or hospitalizations (OR 0.93; 95% CI 0.57-1.51; p=0.771).

**Table 4:** Multivariate analysis of association with any return visit, number of return visits, and and ED visit or hospitalization.

| Characteristic |  | Any return visit |  | Number of return visits |  | ED visit or hospitalization |  |
| --- | --- | --- | --- | --- | --- | --- | --- |
|  |  | Odds ratio (95% CI) | p-value | Rate ratio (95% CI) | p-value | Odds ratio (95% CI) | p-value |
| Intervention | (yes vs no) | 0.79 (0.70, 0.90) | 0.0003 | 0.83 (0.70, 0.97) | 0.0226 | 0.93 (0.57, 1.51) | 0.7708 |
| Age | (<6 months vs 6-23 months) | 1.24 (0.99, 1.56) | 0.0595 | 1.26 (0.99, 1.59) | 0.0602 | 1.05 (0.45, 2.45) | 0.9151 |
|  | (2-4 years vs 6-23 months) | 0.88 (0.71, 1.10) | 0.2645 | 1.01 (0.86, 1.18) | 0.9284 | 1.21 (0.80, 1.83) | 0.3678 |
|  | (5-11 years vs 6-23 months) | 0.68 (0.50, 0.93) | 0.0160 | 0.86 (0.68, 1.10) | 0.2246 | 0.82 (0.45, 1.47) | 0.5004 |
|  | (12-17 years vs 6-23 months) | 1.17 (0.80, 1.71) | 0.4203 | 1.26 (1.01, 1.57) | 0.0376 | 3.00 (1.53, 5.9) | 0.0014 |
| Insurance | (Private vs Public) | 1.20 (0.87, 1.66) | 0.2660 | 1.54 (0.87, 2.74) | 0.1392 | 1.08 (0.75, 1.56) | 0.6940 |
|  | (Other/None vs Public) | 0.63 (0.35, 1.14) | 0.1289 | 1.36 (0.90, 2.05) | 0.1417 | 0.41 (0.18, 0.93) | 0.0319 |
| Illness Duration | (per 7 days) | 1.61 (1.06, 2.47) | 0.0270 | 1.37 (1.12, 1.69) | 0.0022 | 1.55 (1.26, 1.90) | <0.0001 |
| Sex | (Female vs Male) | 0.90 (0.76, 1.06) | 0.2089 | 1.06 (0.91, 1.23) | 0.4559 | 1.01 (0.71, 1.43) | 0.9760 |
| Race | (Black vs White) | 0.49 (0.37, 0.66) | <0.0001 | 0.61 (0.47, 0.78) | <0.0001 | 0.68 (0.45, 1.05) | 0.0836 |
|  | (Other/Unknown vs White) | 0.67 (0.47, 0.95) | 0.0241 | 0.78 (0.62, 1.00) | 0.0471 | 0.82 (0.64, 1.06) | 0.1276 |
| Ethnicity | (Hispanic vs Non-Hispanic)) | 1.15 (0.87, 1.54) | 0.3218 | 1.05 (0.80, 1.37) | 0.7472 | 0.89 (0.55, 1.42) | 0.6171 |
| Diarrhea | (yes vs no) | 1.36 (0.90, 2.05) | 0.1455 | 1.23 (0.87, 1.72) | 0.2411 | 0.75 (0.37, 1.53) | 0.4302 |
| Fever | (yes vs no) | 1.35 (1.02, 1.77) | 0.0347 | 1.32 (1.06, 1.65) | 0.0149 | 1.60 (0.74, 3.44) | 0.2325 |
| Constipation | (yes vs no) | 1.17 (0.67, 2.04) | 0.5907 | 1.02 (0.67, 1.54) | 0.9428 | 1.01 (0.48, 2.14) | 0.9796 |
| Number of Stools | (per 10 stools) | 1.09 (0.96, 1.25) | 0.1797 | 1.07 (0.96, 1.18) | 0.2080 | 1.57 (1.27, 1.93) | <0.0001 |
| Quarter by Time | (Spring 2015 vs Summer 2015) | 0.93 (0.75, 1.15) | 0.4958 | 1.12 (0.95, 1.32) | 0.1691 | 0.55 (0.38, 0.81) | 0.0020 |
|  | (Fall 2015 vs Summer 2015) | 1.10 (0.87, 1.40) | 0.4178 | 1.10 (0.83, 1.44) | 0.5115 | 0.92 (0.56, 1.52) | 0.7504 |
|  | (Winter 2016 vs Summer 2015) | 1.22 (0.88, 1.71) | 0.2380 | 1.75 (1.42, 2.15) | <.0001 | 1.14 (0.77, 1.68) | 0.5228 |
|  | (Spring 2016 vs Summer 2015) | 1.82 (1.32, 2.51) | 0.0003 | 1.35 (1.04, 1.76) | 0.0260 | 1.51 (0.97, 2.36) | 0.0665 |
|  | (Summer 2016 vs Summer 2015) | 2.15 (1.39, 3.33) | 0.0006 | 1.26 (0.94, 1.70) | 0.1210 | 1.09 (0.73, 1.61) | 0.6783 |
| Pathogen Type | (Bacteria vs No Pathogen) | 0.99 (0.49, 1.99) | 0.9723 | 0.97 (0.60, 1.55) | 0.8847 | 1.31 (0.86, 1.99) | 0.2079 |
|  | (Virus vs No Pathogen) | 0.68 (0.42, 1.10) | 0.1172 | 0.81 (0.40, 1.64) | 0.5571 | 0.80 (0.65, 0.99) | 0.0391 |
|  | (Protozoa vs No Pathogen) | 1.40 (0.78, 2.50) | 0.2624 | 0.95 (0.61, 1.49) | 0.8317 | 0.39 (0.08, 2.05) | 0.2667 |
|  | (C diff vs No Pathogen) | 0.83 (0.27, 2.61) | 0.7559 | 1.06 (0.81, 1.38) | 0.6862 | 0.26 (0.05, 1.29) | 0.0987 |
|  | (Missing vs No Pathogen) | 1.02 (0.54, 1.95) | 0.9432 | 0.75 (0.56, 1.00) | 0.0533 | 1.12 (0.69, 1.83) | 0.6392 |

Multivariate sensitivity analyses omitting CMH to account for the *Shigella spp.* outbreak yielded similar results for any return visit (OR 0.74; 95% CI 0.68-0.80; P<0.001; Supplemental Table 1; number of return visits rate ratio 0.77; 95% CI 0.72-0.83; P<0.001). In the sensitivity analysis the odds of return to the ED or hospitalization was reduced during the intervention period (OR 0.70; 95% CI 0.56-0.86; P=0.001).

Results of multivariate analyses of any return visit within subgroups were broadly consistent with the main analysis (**Figure 2**). Analyses restricted to individual sites had substantial uncertainty due to collinearity between intervention periods and time within site. The sensitivity analysis omitting observations from CMH because of the *Shigella spp.* outbreak were similar to the main analysis with slighty greater point estimates of the reduction in rate of return visits. **(Supplemental table 2)** There was no significant difference between the pre- and intervention periods in the proportion of children who received an antibiotic in the ED (3.5% vs 4.1%) or after discharge (4.2% vs 3.8%) in univariate or multivariate analysis. **(Table 3)** However, numerically more patients received the appropriate treatment for a potentially treatable pathogen during the intervention period with multiplex molecular testing; 12 (19.6%) of 61 vs 7 (11.3%) of 62 (P=0.22) and appropriate treatment for *Shigella*; 11 (46%) of 24 vs 7 (21%) of 33 (P = 0.08).

**Figure 2:**
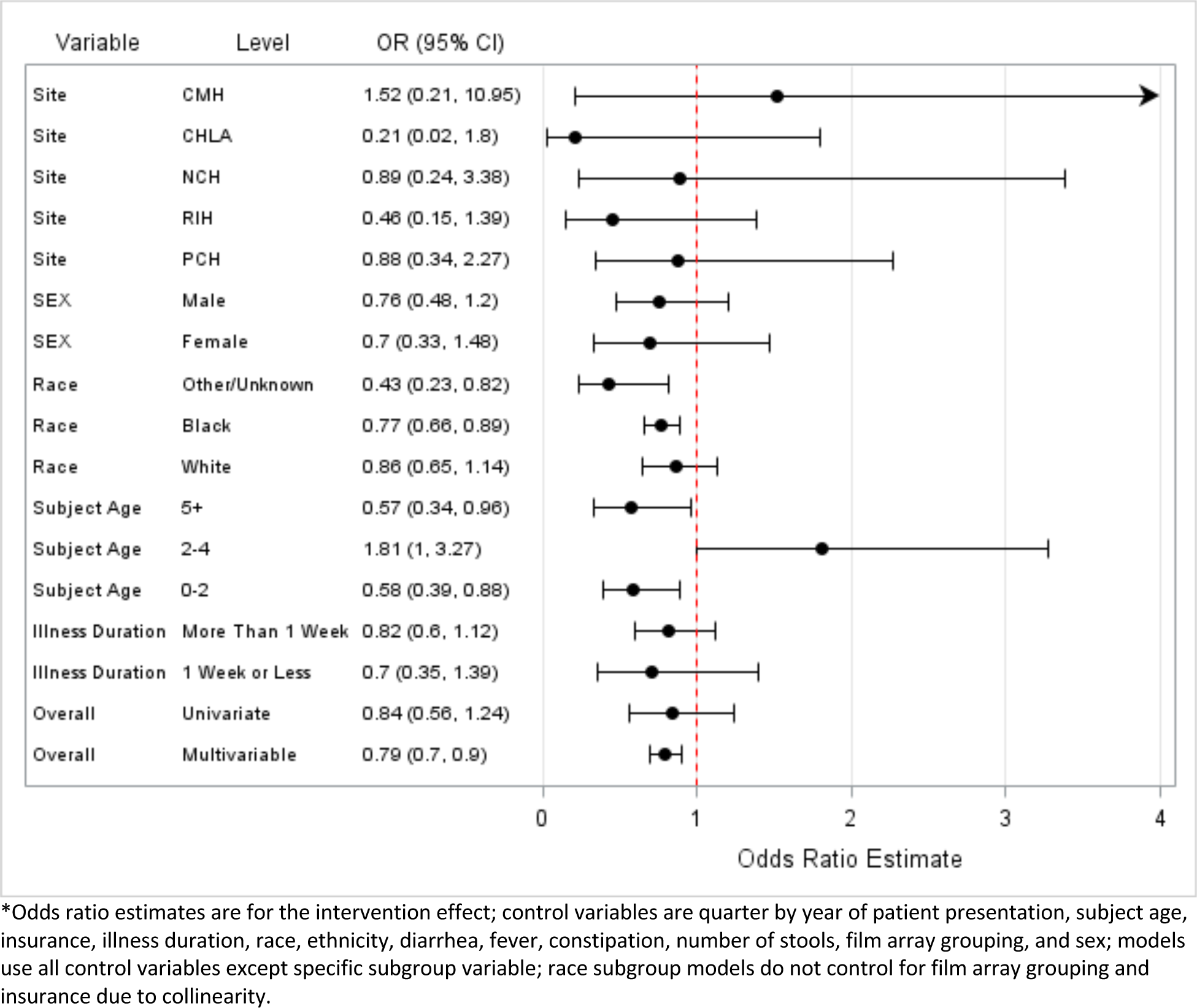
**Forest plot of subgroup analyses for any return visits.\***

Impact of illness on the family was notable. Half of caregivers in both periods reported missing work (median 2 days; IQR 1-3.5). Diarrhea developed in one or more family contacts after the child was enrolled in 30% of cases. These factors did not differ by intervention period. Of 413 caregivers in the intervention arm who reported learning the etiology of their child’s illness in the intervention period, 215 (52%) reported they were “very comfortable” caring for their child’s illness compared to 304 (53%) of caregivers in the pre-intervention period.

## DISCUSSION

Recent technological advances have resulted in the availability of rapid, sensitive, and comprehensive molecular diagnostics for diarrheal illness.[7, 8]. Compared with the wealth of evidence supporting rehydration for pediatric gastroenteritis management, there is a paucity of studies demonstrating impact of these diagnostics on health and patient-centered outcomes, their cost effectiveness, or when they are most useful. [7, 16, 17] In this multi-center prospective pragmatic study, we found that among children presenting to EDs with gastroenteritis, the use of a multiplex molecular diagnostic was associated with a 21% reduction in the likelihood of return visits to healthcare compared with clinician-selected diagnostic testing. As in other studies, we found that multiplex molecular testing substantially increased detection of potential pathogens, including treatable pathogens and those that warranted withholding of empiric antibiotics. Awareness of diagnostic testing results was not associated with increased caregiver comfort in caring for the child’s illness.

We prospectively compared clinician-, patient-, and family-based outcomes before and after introduction of a multiplex molecular diagnostic for children presenting to an ED with gastroenteritis. The pre-intervention and intervention periods were not balanced for a number of key variables associated with diarrheal disease, and our unadjusted analysis showed no differences in likelihood of follow-up visits. After adjusting for factors including demographics, pathogen type, and season, we found a 21% reduction in odds of any return visits to healthcare (OR 0.79; 95% CI 0.70-0.90), our primary endpoint. Stratified analyses suggested the effect was most pronounced in children with a viral pathogen detected (OR 0.56; 95% CI 0.39-0.79), and children enrolled in the winter (OR 0.44; 95% CI 0.23-0.85), the season with the highest rates of viral gastroenteritis.[18] Knowledge of a virus as cause of illness may have reduced clinician and patient uncertainty about the clinical course and need for re-evaluation.

Advantages of molecular diagnostics over traditional stool culture include increased sensitivity for pathogen detection and faster turnaround time, potentially resulting in improvements in appropriate use of antibiotics. We found that multiplex molecular testing of all patients identified a potentially treatable pathogen in a higher proportion (17.3%) of patients during the pre-intervention period compared to clinician-ordered testing (3.2%) as well as clinically relevant pathogens (22% vs 2.8%). We did not detect a significant improvement in overall appropriate antibiotic prescribing among children with a treatable pathogen, although consistent with other studies, fewer than 20% of children had a treatable pathogen in either period.[9, 19, 20] We observed an increase in percentage of patients with *Shigella spp.*/EIEC who received appropriate antibiotics during the intervention period although this was not statistically significant (p = 0.08). Knowledge of the outbreak in the pre-intervention period may have increased appropriate empiric therapy, and therefore reduced the impact of the intervention. Recent observational studies have shown that introduction of a multiplex GI PCR panel was associated with improvements in the selection and duration of antimicrobial therapy [7, 10] [21] [22] [23]

The burden of pediatric gastroenteritis extends beyond the patient, and includes significant nonmedical costs, the largest of which are foregone earnings due to missed work and transportation costs. Half of caregivers reported missing work, and approximately 30% of household contacts subsequently developed diarrhea. Although we did not observe differences in days of work missed with the intervention, it is likely that the decrease in return visits translates to economic benefits. Cost savings attributable to a multiplex diagnostic panel has been shown in an inpatient population [16, 21, 22] and in an a study of adult based on claims data[24], but not to our knowledge among pediatric ED patients. Future studies are warranted to examine the cost impacts and cost-effectiveness.

Further studies are needed to identify the types of patients most likely to benefit from molecular diagnostics, as opposed to the “test all” strategy used in this study. Unfortunately, existing recommendations for selecting patients most likely to have bacterial infection do not perform optimally.[25] Novel strategies for stewardship of molecular diagnostics are needed[26, 27], including the development of clinical decision support tools.[28, 29]

The patterns of organisms detected found in our study is consistent with prior studies using multiplex detection for children with diarrhea in high-income countries. We found EPEC, norovirus, and *C. difficile* to be the most commonly detected pathogens overall, similar to previous studies.[9, 20] However, interpreting the results of molecular detection of potential pathogens can be challenging, since not all molecular detections correspond to the etiology of symptoms due to detection of asymptomatic carriage or lack of specificity of the target.[30] This is particularly problematic for EPEC and *C. difficile*. We did not observe increased prescription of drugs for *C. difficile*, despite increased detection. EPEC was commonly detected (>15% of samples), but the clinical interpretation of detection of the single gene target (*eae*) is unknown. Case-control studies using molecular diagnostics, such as ones already conducted in other settings [31] [32] are urgently needed in the US to determine the relevance of organisms detected through various gene targets. Quantitative PCR thresholds from case-control studies may increase specificity. [33]

Our study has limitations. First, despite the stepped-wedge design of the enrollment across sites, none of the sites were able to capture the same months during the pre- and post-intervention period. Thus, our analysis required adjustment by season and time of year. Second, a *Shigella* outbreak occurred at one site during the study[15], which led to a higher proportion of *Shigella* detected than would be expected. However, our sensitivity analyses excluding this site confirmed the results. Third, only 24% of children were able to get results before discharge from the ED because of failure to produce a specimen. Use of rectal swabs might improve this and increase the impact of diagnosis.[34] Fourth, power was modest. Lastly, a larger proportion (15% vs. 4%) of those enrolled in the pre-intervention phase were lost to follow-up compared to the intervention phase.

In conclusion, in a multi-center prospective pragmatic study of a multi-pathogen molecular diagnostic panel for pediatric gastroenteritis, we found that use of panel improved pathogen detection and decreased likelihood of return visits to healthcare. Further research is needed to identify patients most likely to benefit from the use of these tests and to optimize the cost effectiveness of different diagnostic approaches to pediatric gastroenteritis.

## Data Availability

Availability of the data from this study will be determined after publication

## ACKNOWLEDGEMENTS

We thank Julian Dorsch, Tania Baca, and the ED physicians, pharmacists and research coordinators who made this study possible. We are indebted to the families who participated.

---

**Supplemental Table 1:** Pathogens detected by multiplex PCR and those considered “treatable” or “clinically actionable,” defined as either treatable or the use of antibiotics would be potentially harmful

| Pathogen | Treatable | Clinically actionable |
| --- | --- | --- |
| **Bacteria** |  |  |
| *Campylobacter spp. (C. jejuni, C. coli, C. upsaliensis)* | X | X |
| *C. difficile* toxin A/B |  |  |
| *Plesiomonas shigelloides* | X | X |
| *Salmonella spp.* |  | X |
| *Yersinia enterocolitica,* | X | X |
| *Vibrio spp. (V. parahaemolyticus, V. vulnificus, V. cholerae)* | X | X |
| Enteraggregative *E. coli* [EAEC] |  |  |
| Enteropathogenic *E. coli* [EPEC] |  |  |
| Enterotoxigenic *E. coli* [ETEC] |  |  |
| Shigatoxin-producing *E. coli* [STEC] |  | X |
| *Shigella spp./*Enteroinvasive *E. coli* [EIEC] | X | X |
| **Viruses** |  |  |
| adenovirus F 40/41 |  |  |
| astrovirus |  |  |
| norovirus GI/GII |  |  |
| rotavirus A |  |  |
| sapovirus |  |  |
| **Protozoa** |  |  |
| *Cryptosporidium spp.* | X | X |
| *Cyclospora cayatensi* | X | X |
| *Entamoeba histolytica* | X | X |
| *Giardia lamblia* | X | X |

**Supplemental Table 2:** Sensitivity multivariate analysis results for return visit endpoints without CMH observations.

| Characteristic |  | Any return visit |  | Number of return visits |  | ED visit or hospitalization |  |
| --- | --- | --- | --- | --- | --- | --- | --- |
|  |  | Odds ratio (95% CI) | p-value | Rate ratio (95% CI) | p-value | Odds ratio (95% CI) | p-value |
| Intervention | (yes vs no) | 0.74 (0.68, 0.80) | <0.0001 | 0.77 (0.72, 0.83) | <0.0001 | 0.70 (0.56, 0.86) | 0.0010 |
| Age | (<6 months vs 6-23 months) | 1.11 (1.00, 1.23) | 0.0524 | 1.37 (1.15, 1.63) | 0.0005 | 0.78 (0.35, 1.73) | 0.5338 |
|  | (2-4 years vs 6-23 months) | 0.88 (0.69, 1.14) | 0.3353 | 1.02 (0.85, 1.22) | 0.8277 | 1.04 (0.64, 1.69) | 0.8836 |
|  | (5-11 years vs 6-23 months) | 0.74 (0.52, 1.05) | 0.0907 | 0.95 (0.80, 1.13) | 0.5737 | 0.99 (0.59, 1.68) | 0.9732 |
|  | (12-17 years vs 6-23 months) | 1.35 (1.00, 1.81) | 0.0477 | 1.17 (1.00, 1.36) | 0.0490 | 3.60 (1.81, 7.14) | 0.0003 |
| Insurance | (Private vs Public) | 1.22 (0.86, 1.73) | 0.2706 | 1.47 (0.74, 2.92) | 0.2754 | 1.07 (0.69, 1.67) | 0.7666 |
|  | (Other/None vs Public) | 0.70 (0.34, 1.45) | 0.3396 | 1.30 (0.78, 2.17) | 0.3094 | 0.33 (0.06, 1.84) | 0.2058 |
| Illness Duration | (per 7 days) | 1.44 (0.97, 2.14) | 0.0706 | 1.26 (1.08, 1.48) | 0.0035 | 1.41 (1.31, 1.52) | <0.0001 |
| Sex | (Female vs Male) | 0.91 (0.74, 1.11) | 0.3429 | 1.04 (0.89, 1.22) | 0.6183 | 1.07 (0.73, 1.56) | 0.7310 |
| Race | (Black vs White) | 0.45 (0.34, 0.58) | <0.0001 | 0.56 (0.46, 0.70) | <0.0001 | 0.53 (0.35, 0.79) | 0.0020 |
|  | (Other/Unknown vs White) | 0.76 (0.60, 0.96) | 0.0219 | 0.85 (0.71, 1.02) | 0.0793 | 0.96 (0.85, 1.08) | 0.4984 |
| Ethnicity | (Hispanic vs Non-Hispanic) | 1.11 (0.80, 1.54) | 0.5274 | 1.01 (0.75, 1.36) | 0.9508 | 0.93 (0.52, 1.67) | 0.8129 |
| Diarrhea | (yes vs no) | 1.47 (0.97, 2.23) | 0.0710 | 1.27 (0.86, 1.89) | 0.2292 | 1.12 (0.70, 1.79) | 0.6350 |
| Fever | (yes vs no) | 1.28 (0.94, 1.73) | 0.1141 | 1.28 (1.00, 1.64) | 0.0481 | 1.69 (0.69, 4.16) | 0.2550 |
| Constipation | (yes vs no) | 0.95 (0.63, 1.43) | 0.8099 | 0.90 (0.64, 1.26) | 0.5258 | 0.90 (0.41, 1.98) | 0.7860 |
| Number of Stools | (per 10 stools) | 1.09 (0.93, 1.27) | 0.2910 | 1.06 (0.94, 1.19) | 0.3176 | 1.53 (1.23, 1.91) | 0.0002 |
| Quarter by Time | (Spring 2015 vs Summer 2015) | 0.80 (0.65, 0.97) | 0.0261 | 1.02 (0.89, 1.16) | 0.8248 | 0.45 (0.35, 0.59) | <.0001 |
|  | (Fall 2015 vs Summer 2015) | 0.97 (0.71, 1.32) | 0.8272 | 0.97 (0.76, 1.25) | 0.8304 | 0.74 (0.47, 1.19) | 0.2184 |
|  | (Winter 2016 vs Summer 2015) | 1.14 (0.95, 1.36) | 0.1556 | 1.63 (1.43, 1.86) | <.0001 | 1.07 (0.72, 1.59) | 0.7496 |
|  | (Spring 2016 vs Summer 2015) | 1.78 (1.50, 2.12) | <.0001 | 1.19 (1.11, 1.28) | <.0001 | 1.28 (0.89, 1.85) | 0.1803 |
|  | (Summer 2016 vs Summer 2015) | 1.72 (1.69, 1.76) | <.0001 | 1.21 (1.03, 1.42) | 0.0215 | 0.93 (0.61, 1.43) | 0.7488 |
| Pathogen Type | (Bacteria vs No Pathogen) | 0.98 (0.37, 2.56) | 0.9623 | 0.92 (0.51, 1.66) | 0.7798 | -* | - |
|  | (Virus vs No Pathogen) | 0.60 (0.36, 0.98) | 0.0419 | 0.57 (0.24, 1.36) | 0.2042 | - | - |
|  | (Protozoa vs No Pathogen) | 1.47 (0.59, 3.64) | 0.4089 | 0.81 (0.60, 1.09) | 0.1645 | - | - |
|  | (C diff vs No Pathogen) | 0.55 (0.12, 2.58) | 0.4471 | 1.08 (0.77, 1.53) | 0.6463 | - | - |
|  | (Missing vs No Pathogen) | 0.80 (0.48, 1.35) | 0.4088 | 0.70 (0.53, 0.92) | 0.0099 | - | - |

\*Pathogen type was removed due to model instability.

**Supplemental Figure 1:**
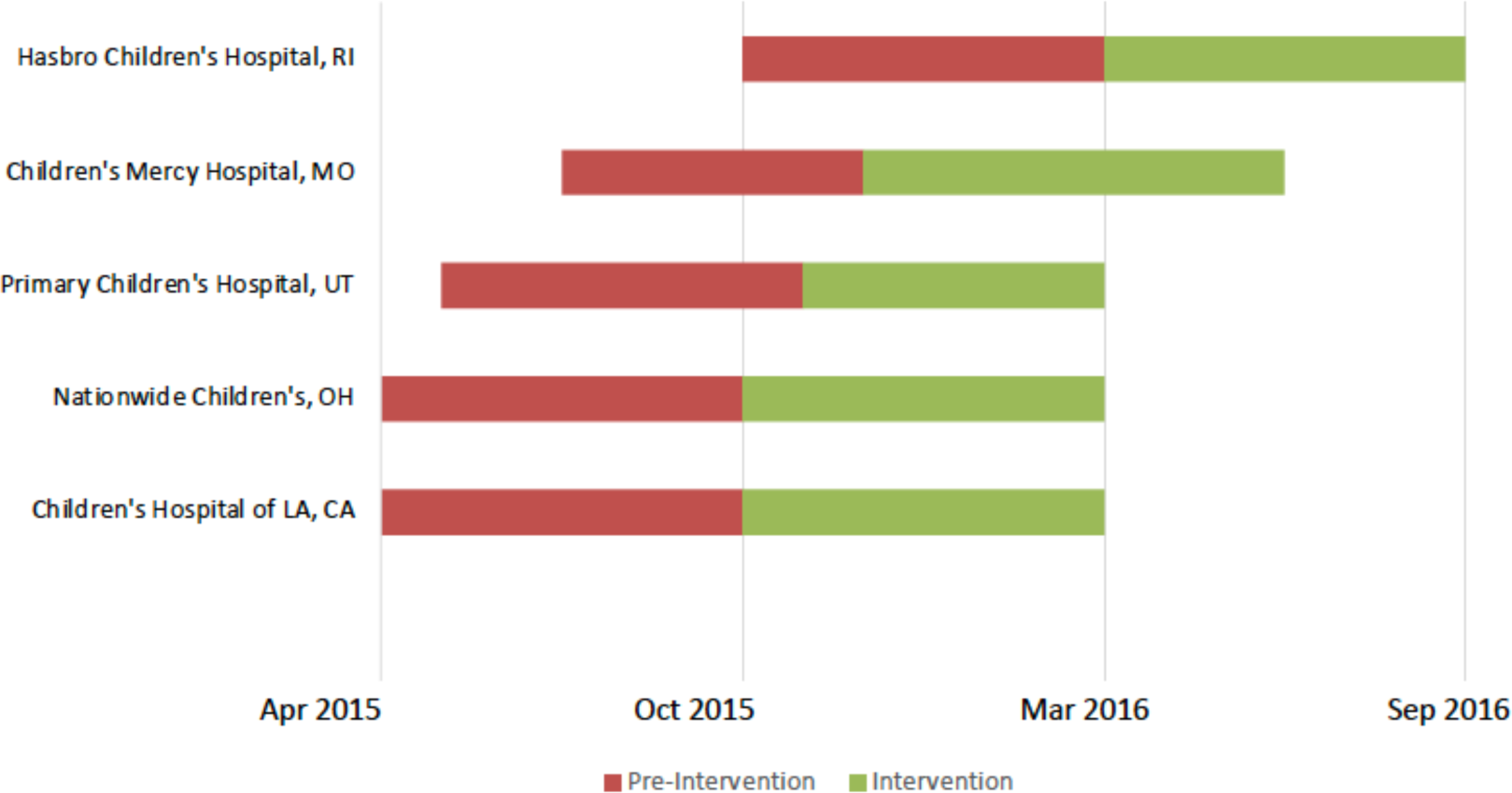
Time course of enrollment in Pre-intervention and Intervention periods by study site

## REFERENCES

1. Jones TF, McMillian MB, Scallan E, et al. A population-based estimate of the substantial burden of diarrhoeal disease in the United States; FoodNet, 1996-2003. Epidemiol Infect 2007; 135(2): 293–301.

2. G. B. D. Diarrhoeal Disease Collaborators. Estimates of the global, regional, and national morbidity, mortality, and aetiologies of diarrhoea in 195 countries: a systematic analysis for the Global Burden of Disease Study 2016. Lancet Infect Dis 2018; 18(11): 1211–28.

3. Shane AL, Mody RK, Crump JA, et al. 2017 Infectious Diseases Society of America Clinical Practice Guidelines for the Diagnosis and Management of Infectious Diarrhea. Clin Infect Dis 2017; 65(12): e45–e80.

4. Guarino A, Ashkenazi S, Gendrel D, et al. European Society for Pediatric Gastroenterology, Hepatology, and Nutrition/European Society for Pediatric Infectious Diseases evidence-based guidelines for the management of acute gastroenteritis in children in Europe: update 2014. J Pediatr Gastroenterol Nutr 2014; 59(1): 132–52.

5. Buss SN, Leber A, Chapin K, et al. Multicenter evaluation of the BioFire FilmArray gastrointestinal panel for etiologic diagnosis of infectious gastroenteritis. J Clin Microbiol 2015; 53(3): 915–25.

6. Binnicker MJ. Multiplex Molecular Panels for Diagnosis of Gastrointestinal Infection: Performance, Result Interpretation, and Cost-Effectiveness. J Clin Microbiol 2015; 53(12): 3723–8.

7. Cybulski RJ, Jr., Bateman AC, Bourassa L, et al. Clinical impact of a Multiplex Gastrointestinal PCR Panel in Patients with Acute Gastroenteritis. Clin Infect Dis 2018.

8. Khare R, Espy MJ, Cebelinski E, et al. Comparative evaluation of two commercial multiplex panels for detection of gastrointestinal pathogens by use of clinical stool specimens. J Clin Microbiol 2014; 52(10): 3667–73.

9. Stockmann C, Pavia AT, Graham B, et al. Detection of 23 Gastrointestinal Pathogens Among Children Who Present With Diarrhea. J Pediatric Infect Dis Soc 2017; 6(3): 231–8.

10. Torres-Miranda D, Akselrod H, Karsner R, et al. Use of BioFire FilmArray gastrointestinal PCR panel associated with reductions in antibiotic use, time to optimal antibiotics, and length of stay. BMC Gastroenterol 2020; 20(1): 246.

11. Fang FC, Patel R. 2017 Infectious Diseases Society of America Infectious Diarrhea Guidelines: A View From the Clinical Laboratory. Clin Infect Dis 2017; 65(12): 1974–6.

12. Marder EP, Cieslak PR, Cronquist AB, et al. Incidence and Trends of Infections with Pathogens Transmitted Commonly Through Food and the Effect of Increasing Use of Culture-Independent Diagnostic Tests on Surveillance - Foodborne Diseases Active Surveillance Network, 10 U.S. Sites, 2013-2016. MMWR Morb Mortal Wkly Rep 2017; 66(15): 397–403.

13. Freedman DA. On The So-Called “Huber Sandwich Estimator” and “Robust Standard Errors”. The American Statistician 2006; 60(4): 299–302.

14. Zeger SL, Liang KY. Longitudinal data analysis for discrete and continuous outcomes. Biometrics 1986; 42(1): 121–30.

15. Kanwar N, Jackson J, Bardsley T, et al. Impact of Rapid Molecular Multiplex Gastrointestinal Pathogen Testing in Management of Children during a Shigella Outbreak. J Clin Microbiol 2023; 61(3): e0165222.

16. Beal SG, Tremblay EE, Toffel S, Velez L, Rand KH. A Gastrointestinal PCR Panel Improves Clinical Management and Lowers Health Care Costs. J Clin Microbiol 2018; 56(1).

17. Freeman K, Mistry H, Tsertsvadze A, et al. Multiplex tests to identify gastrointestinal bacteria, viruses and parasites in people with suspected infectious gastroenteritis: a systematic review and economic analysis. Health technology assessment (Winchester, England) 2017; 21(23): 1–188.

18. Payne DC, Parashar UD. Rapid advances in understanding viral gastroenteritis from domestic surveillance. Emerg Infect Dis 2013; 19(8): 1189.

19. Denno DM, Shaikh N, Stapp JR, et al. Diarrhea etiology in a pediatric emergency department: a case control study. Clin Infect Dis 2012; 55(7): 897–904.

20. Nicholson MR, Van Horn GT, Tang YW, et al. Using Multiplex Molecular Testing to Determine the Etiology of Acute Gastroenteritis in Children. J Pediatr 2016; 176: 50–6 e2.

21. Cotter JM, Thomas J, Birkholz M, Ambroggio L, Holstein J, Dominguez SR. Clinical Impact of a Diagnostic Gastrointestinal Panel in Children. Pediatrics 2021; 147(5).

22. Axelrad JE, Freedberg DE, Whittier S, Greendyke W, Lebwohl B, Green DA. Impact of Gastrointestinal Panel Implementation on Health Care Utilization and Outcomes. J Clin Microbiol 2019; 57(3).

23. Brendish NJ, Beard KR, Malachira AK, et al. Clinical impact of syndromic molecular point-of-care testing for gastrointestinal pathogens in adults hospitalised with suspected gastroenteritis (GastroPOC): a pragmatic, open-label, randomised controlled trial. Lancet Infect Dis 2023.

24. Moon RC, Bleak TC, Rosenthal NA, et al. Relationship between Diagnostic Method and Pathogen Detection, Healthcare Resource Use, and Cost in U.S. Adult Outpatients Treated for Acute Infectious Gastroenteritis. J Clin Microbiol 2023; 61(2): e0162822.

25. Tarr GAM, Chui L, Lee BE, et al. Performance of Stool-testing Recommendations for Acute Gastroenteritis When Used to Identify Children With 9 Potential Bacterial Enteropathogens. Clin Infect Dis 2019; 69(7): 1173–82.

26. Messacar K, Parker SK, Todd JK, Dominguez SR. Implementation of Rapid Molecular Infectious Disease Diagnostics: the Role of Diagnostic and Antimicrobial Stewardship. J Clin Microbiol 2017; 55(3): 715–23.

27. Tarr GAM, Tarr PI. Pediatric Enteric Diagnostic Stewardship: The Right Test in the Right Context. Pediatrics 2021; 147(5).

28. Nelson EJ, Khan AI, Keita AM, et al. Improving Antibiotic Stewardship for Diarrheal Disease With Probability-Based Electronic Clinical Decision Support: A Randomized Crossover Trial. JAMA Pediatr 2022; 176(10): 973–9.

29. Brintz BJ, Howard JI, Haaland B, et al. Clinical predictors for etiology of acute diarrhea in children in resource-limited settings. PLoS Negl Trop Dis 2020; 14(10): e0008677.

30. Tarr GAM, Tarr PI, Freedman SB. Clinical interpretation of enteric molecular diagnostic tests. Clin Microbiol Infect 2019; 25(12): 1454–6.

31. Kotloff KL, Nataro JP, Blackwelder WC, et al. Burden and aetiology of diarrhoeal disease in infants and young children in developing countries (the Global Enteric Multicenter Study, GEMS): a prospective, case-control study. Lancet 2013; 382(9888): 209–22.

32. Alsuwaidi AR, Al Dhaheri K, Al Hamad S, et al. Etiology of diarrhea by multiplex polymerase chain reaction among young children in the United Arab Emirates: a case-control study. BMC Infect Dis 2021; 21(1): 7.

33. Liu J, Platts-Mills JA, Juma J, et al. Use of quantitative molecular diagnostic methods to identify causes of diarrhoea in children: a reanalysis of the GEMS case-control study. Lancet 2016; 388(10051): 1291–301.

34. Freedman SB, Xie J, Nettel-Aguirre A, et al. Enteropathogen detection in children with diarrhoea, or vomiting, or both, comparing rectal flocked swabs with stool specimens: an outpatient cohort study. Lancet Gastroenterol Hepatol 2017; 2(9): 662–9.

